# Effective Natural Language Processing Algorithms for Gout Flare Early Alert from Chief Complaints

**DOI:** 10.1101/2023.11.28.23299150

**Authors:** Lucas Lopes Oliveira, Xiaorui Jiang, Aryalakshmi Nellippillipathil Babu, Poonam Karajagi, Alireza Daneshkhah

## Abstract

Early identification of acute gout is crucial, enabling healthcare professionals to implement targeted interventions for rapid pain relief and preventing disease progression, ensuring improved long-term joint function. In this study, we comprehensively explored the potential of gout flare (GF) early detection based on nurse chief complaint notes in the Emergency Department (ED). Addressing the challenge of identifying GFs prospectively during an ED visit, where documentation is typically minimal, our research focuses on employing alternative Natural Language Processing (NLP) techniques to enhance the detection accuracy. We investigate GF detection algorithms using both sparse representations by traditional NLP methods and dense encodings by medical domain-specific Large Language Models (LLMs), distinguishing between generative and discriminative models. Three methods are used to alleviate the issue of severe data imbalance, including oversampling, class weights, and focal loss. Extensive empirical studies are done on the Gout Emergency Department Chief Complaint Corpora. Sparse text representations like tf-idf proved to produce strong performance, achieving higher than 0.75 F1 Score. The best deep learning models are RoBERTa-Large-PM-M3-Voc and BioGPT, with the best F1 Scores on each dataset with a 0.8 on the 2019 dataset and a 0.85 F1 Score the 2020 dataset. We concluded that although discriminative LLMs performed better for this classification task, compared to generative LLMs, a combination of using generative models as feature extractors and employing support vector machine for classification yields promising results comparable to those obtained with discriminative models.

## 1. Introduction

More than 9 million Americans suffer from gout [1], which is the most prevalent type of inflammatory arthritis among men, affecting over 5% of them. According to the U.S. National Emergency Department Sample (NEDS), gout accounts for more than 200,000 visits to the Emergency Department (ED) every year, making up 0.2% of all ED visits and costing more than $280 million in annual charges [2]. It is important to improve the continuity of care for gout patients, especially after an ED visit. Often, gout flares (GF) treated in the ED lack optimal follow-up care, necessitating the development of methods for identifying and referring patients with GFs during an ED visit [3]. While retrospective studies have leveraged NLP for GF detection, the prospective identification of patients in real time ED settings presents a unique challenge, especially within the constraints of Emergency Department (ED) environments.

Despite of the success of natural language processing (NLP) techniques in healthcare [4], NLP-based Gout Flare Early Detection (GFED) is in severe lack of study. Only a few were identified, like Zheng et al [5], which however worked on Electronic Medical Records. The problem of early warning of acute GFs becomes more challenging in the ED setting where only chief complaints of patients are taken by nurses in an extremely succinct format. It is of paramount challenge to develop an effective GFED algorithm using such limited amount of information. The current study tries to address this critical gap by advancing the methodologies proposed by Osborne et al [3]. Our study builds upon the groundwork laid by Osborne et al., who annotated two corpora of ED chief complaint notes for GFs and paves the way for our exploration of effective text representation methods and state-of-the-art medical/clinical Large Language Models (LLM).

### 1.1. Rationale for Using Large Language Models

Large language models, such as BERT [6] (Bidirectional Encoder Representations from Transformers), [7] (Generative Pre-trained Transformer 3), and their variants, have demonstrated remarkable success in a wide range of natural language processing tasks. The use of large language models in text classification offers several compelling reasons:

#### Contextual Understanding

Large language models leverage deep learning techniques to encode contextual information and relationships between words in a sentence. This contextual understanding allows them to capture subtle nuances and semantics, which is especially relevant in the medical domain where precise interpretation of clinical text is vital.

#### Transfer Learning

Pre-training on vast corpora of textual data enables large language models to learn general language patterns. This pre-trained knowledge can be fine-tuned on domain-specific datasets, making them adaptable and effective for text classification tasks in the medical field with relatively limited labelled data.

These technologies have the potential to revolutionize the healthcare industry by enhancing medical decision-making, patient care, and biomedical research. Some tasks in NLP could be automated using LLM such as text classification [8-9], keyword Extraction [10-11], machine translation [12], and text summarization [13]. Furthermore, NLP and LLM can assist in the early detection and diagnosis of diseases by sifting through vast datasets to identify patterns, symptoms, and risk factors.

### 1.2. Gaps and Limitations of Current Literature

While some studies have compared a single generative LLM (GPT) with discriminative LLMs, a comprehensive comparison between multiple domain-specific generative LLMs and discriminative LLMs for disease detection is lacking. Such comparisons are essential to determine the performance disparities between different LLM types and guide the selection of the most suitable model for our specific medical intent classification task.

In light of these gaps, our research aims to bridge these deficiencies in the current literature. We specifically focus on GFED by leveraging domain-specific generative LLMs as feature extractors. Additionally, our study includes comparative analyses of multiple domain specific generative LLMs and discriminative LLMs to gain comprehensive insights into their performance on this particular medical classification task.

### 1.3. Our contributions

In this paper, we make three contributions to the task of gout flare detection from nurse chief complaints. First, we compare the performance of domain specific discriminative and generative models that are fine-tuned for the task. Second, we propose an alternative approach that uses domain specific generative LLMs as feature extractors and support vector machine as classifier. Third, we benchmark our methods against a baseline that uses sparse text representation (tf-idf). Our results demonstrate the effectiveness of using LLMs, such as RoBERTa-Large-PM-M3-Voc, BioELECTRA, and BioGPT, for processing medical text and detecting GFs.

## 2. Materials and Methods

### 2.1. Data Collection

We utilized the dataset of ED chief complaint notes which were annotated by Osborne et al. for the presence of GFs [14]. Each CC text in the dataset was annotated to determine its indication of a GF, a non-GF, or remained unknown in terms of the status of GF. Following this, a manual chart review was conducted by a rheumatologist and a post-doctoral fellow to ascertain the GF status for a small portion of the ED counters. These were served as the gold standard annotations of the real GF status. The corpora contain two datasets for the year 2019 and 202, namely GOUT-CC-2019-CORPUS and GOUT-CC-2020-CORPUS respectively. Table 1 shows the annotation statistics of the two datasets (from Osborne et al. [3]), while Table 2 illustrates some examples. In out experiments, we used the human-annotated samples using Chart Review, as what Osborne et al. did.

**Table 1:** Annotation Statistics of the Gout Flare Chief Complaint Datasets (Osborne et al. [3])

| Dataset Name | GF-POS<br>(Positive) | GF-NEG<br>(Negative) | GF-UNK<br>(Unknown) | Review | Agreement | Cohen's $\kappa$ |
| --- | --- | --- | --- | --- | --- | --- |
| GOUT-CC-2019-CORPUS | 93 | 194 | 13 | CC | 0.883 | 0.825 |
| GOUT-CC-2019-CORPUS* | 70 | 118 | 9 | Chart | 0.849 | 0.774 |
| GOUT-CC-2020-CORPUS | 14 | 7992 | 129 | CC | 0.977 | 0.965 |
| GOUT-CC-2020-CORPUS* | 25 | 232 | 7 | Chart | 0.904 | 0.856 |

\* Used for experiments as Osborne et al. [3]

**Table 2:**
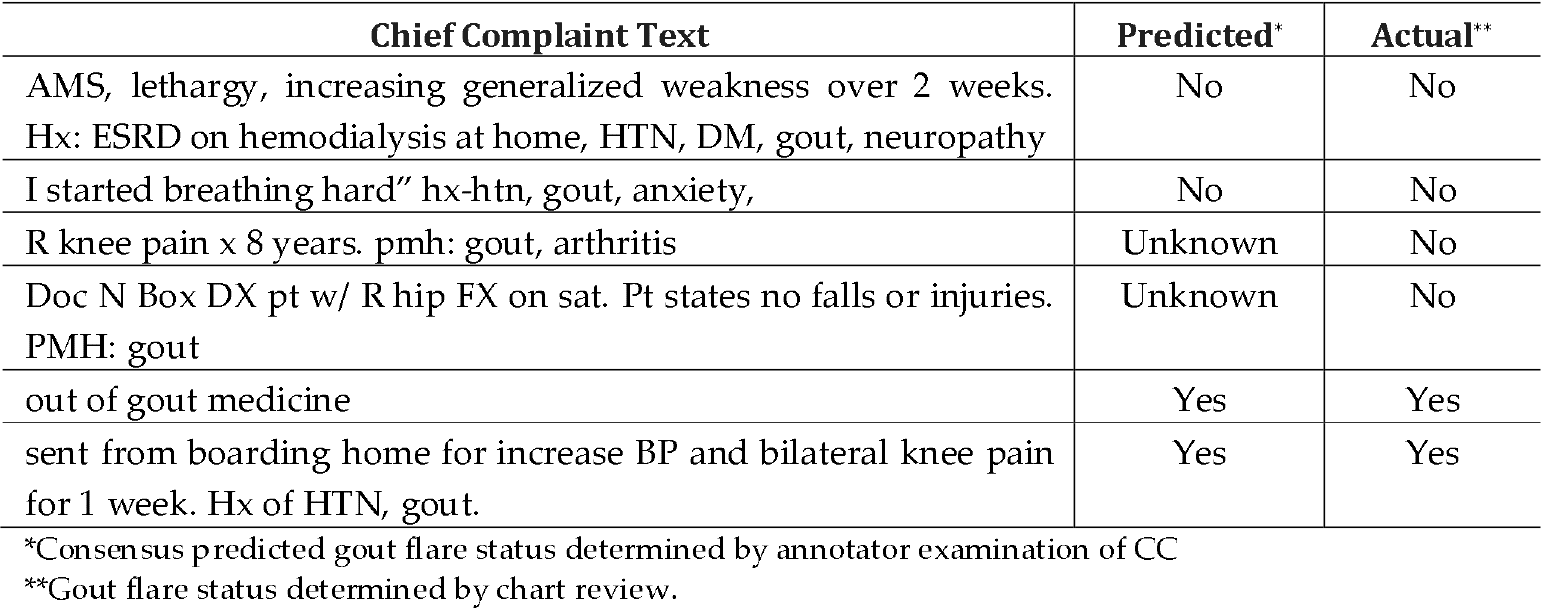
Examples of Chief Complaint Notes for Gout Flare (Osborne et al. [3])

| Chief Complaint Text | Predicted* | Actual** |
| --- | --- | --- |
| AMS, lethargy, increasing generalized weakness over 2 weeks.<br>Hx: ESRD on hemodialysis at home, HTN, DM, gout, neuropathy | No | No |
| I started breathing hard" hx-htn, gout, anxiety, | No | No |
| R knee pain x 8 years. pmh: gout, arthritis | Unknown | No |
| Doc N Box DX pt w/ R hip FX on sat. Pt states no falls or injuries.<br>PMH: gout | Unknown | No |
| out of gout medicine | Yes | Yes |
| sent from boarding home for increase BP and bilateral knee pain<br>for 1 week. Hx of HTN, gout. | Yes | Yes |

\*Consensus predicted gout flare status determined by annotator examination of CC
\*\*Gout flare status determined by chart review.

### 2.2. Feature Extraction

In the feature engineering approach, we extracted the n-grams (n = 1, 2, 3) and tested different combinations of *n*-grams and different feature sizes. CC texts were converted into sparse representations using *tf-idf* (Term Frequency-Inverse Document Frequency) [15] as initial feature values. A linear support vector classifier (Linear SVC) was trained. All implementations were done using the scikit-learn library^1^.

It was hard to extract more advanced syntactic or semantic features due to the noisiness of CC texts. As can be observed from Table 2, CC texts are extremely succinct, often containing a sequence of medical terms or abbreviations, which record the facts reported by patients. Such CCs are not meaningful sentences for us to extract features from the syntactic analysis results. Semantic analysis tools are either immature or non-existent in this particular area. However, we could still observe quite good performances from fine-tuning a machine learning model using the right sparse feature representation of CC texts.

### 2.3. Large Language Models

We employed several LLMs tailored for the medical domain, for their ability to capture intricate patterns within medical text, making them well-suited for discerning nuances in chief complaints related to GF. All LLMs belong to the Transformers family [16] because we hoped that the multi-headed self-attention mechanism of the Transformers architecture could be able to learn the meaningful association between certain words of CC texts to indicate the existence of GF.

#### 2.3.1. Discriminative models

We strategically incorporated three robust discriminative LLMs renowned for their discriminative power—RoBERTa-PM-M3-Vo^2^, BioELECTRA^3^ [17], and BioBART^4^ [18]. These are the domain-specific versions of the RoBERTa [19], Electra [20] and BART [21] models respectively. Although BART was a language model pretrained in a sequence-to-sequence fashion, it can be used equally well and in the same way as a discriminative model [21]. So, we treated it as one representative of the discriminative category. The details of the discriminative LLMs are shown in Table 3.

**Table 3:**
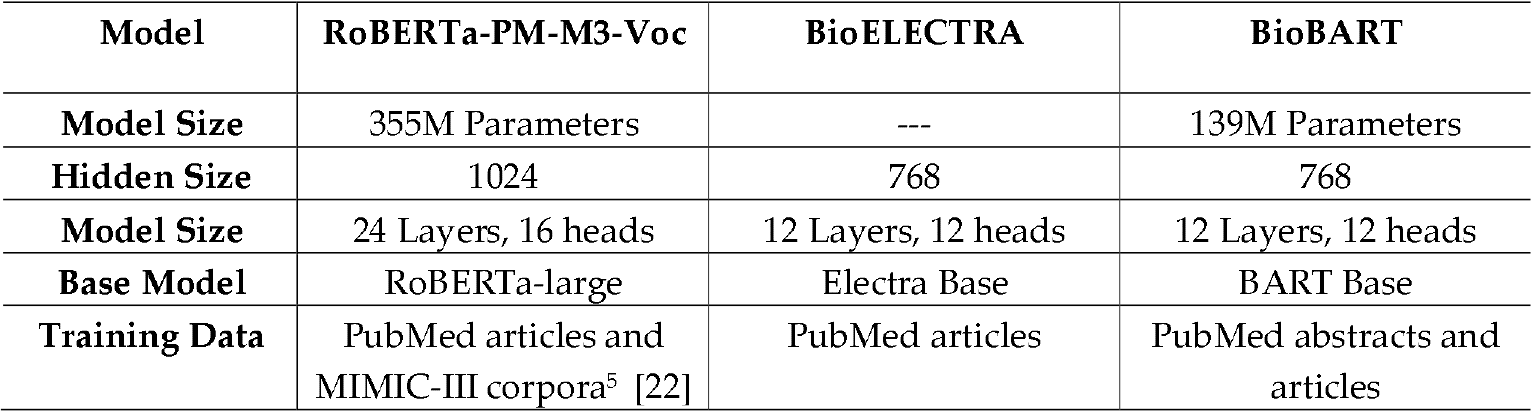
Description of Discriminative LLMs Implemented.

| Model | RoBERTa-PM-M3-Voc | BioELECTRA | BioBART |
| --- | --- | --- | --- |
| Model Size | 355M Parameters | --- | 139M Parameters |
| Hidden Size | 1024 | 768 | 768 |
| Model Size | 24 Layers, 16 heads | 12 Layers, 12 heads | 12 Layers, 12 heads |
| Base Model | RoBERTa-large | Electra Base | BART Base |
| Training Data | PubMed articles and MIMIC-III corpora<sup>5</sup> [22] | PubMed articles | PubMed abstracts and articles |

#### 2.3.2. Generative models

In the realm of generative LLMs, we strategically chose BioGPT^6^ [23], BioMedLM^7^, and PMC_LLaMA_7B^8^ [24] for their renowned scale and exceptional performance in natural language processing tasks. BioGPT and PMC_LLaMA_7B are the domain-specific versions of the GPT-2 [25] and LLaMA [26-27] models respectively, while BioMedLM is a bespoke LLM pretrained for medical applications. These models represent the forefront of generative language understanding, and their comprehensive specifications, training data, and architectural features are elucidated in Table 4.

**Table 4:** Description of Generative LLMs Implemented.

| Model | BioGPT | BioMedLM | PMC_LLaMA_7B |
| --- | --- | --- | --- |
| Model Size | 347M Parameters | 2.7B Parameters | 7B Parameters |
| Hidden Size | 1024 | 2560 | 4096 |
| Model Size | 24 Layers, 16 heads | 32 Layers, 20 heads | 32 Layers, 32 heads |
| Base Model | GPT2-medium | GPT2 | LLaMA_7B |
| Training Data | 15M PubMed abstracts from scratch | All PubMed abstracts and full texts from The Pile benchmark [28]. | 4.8 million Biomedical publications from the S2ORC dataset [29]. |

### 2.4. Fine-tuning

Fine tuning was implemented to improve the models’ ability to understand and capture the nuances in the texts. For the discriminative models full fine tuning was implemented, but for the generative models due to the size of the models and hardware constraints full fine tuning was not possible.

#### 2.4.1. Fine-tuning of Discriminative LLMs

All three discriminative LLMs use a bidirectional encoder as BERT [6]. The encoder part of these models was used to encode each CC text, and the “[CLS]” token was used as the dense representation. For RoBERTa-PM-M3-Voc and BioELECTRA, a further feature transformation was applied. Essentially, the classification head was a Multiple Layer Perceptron (MLP), the hidden layer of which made a nonlinear transformation (of the same size). On the contrary, BioBART used a linear classification head following the tradition of BART usage.

In the fine-tuning process, the following hyperparameters were used: learning rate = 1e-5, epoch number = 10, batch size = 14, early stopping patience = 3. The AdamW optimiser was used for training [30].

#### 2.4.2. Fine-tuning of Generative LLMs

Similarly, generative LLMs were used for encoding CC texts, and the “Extract” token (for all three models as they all belong to the GPT family) were used to extract the dense representation, which was then sent to a linear classification head. Due to their large sizes, the generative LLMs were not fully fine-tuned. Instead, we used LoRA (Low Rank Adaptation) to efficiently adapt LLMs to specific tasks by only modifying a small portion of the whole parameter space.

The main idea behind LoRA is to exploit the low-rank structure of the model’s weight matrices during task adaptation, resulting in reduced memory usage and computational complexity [31]. The idea was inspired by Aghajanyan et al.’s finding that pre-trained language models have a low “intrinsic dimension” meaning that they can still lean efficiently when their weight matrices are randomly projected to a smaller subspace [32].

More precisely, LoRA hypothesizes that updates to model’s weight matrix, *W*_0_, can be represented by a low-rank decomposition, which is given by *W*_0_ + Δ*W* = *W*_0_ + *BA*, where *B* ∈ *R*^*d* × *r*^,*A* ∈ *R*^*r* × *k*^, and *ΔW* = *BA* represents weight updates. During training (i.e., fine-tuning), *W*_0_ is frozen while *A* and *B* contain the trainable parameters.

In our fine-tuning process, we applied the following LoRA parameters:

1. The rank (*r*) of *A* and *B* was set to 8.
2. The LoRA regularization coefficient *α* was set to 16.
3. To prevent overfitting and enhancing model generalisation, we applied a LoRA dropout rate of 0.1.
4. A learning rate of 3e-4 was used, enabling efficient convergence during training.

**Figure 1.**
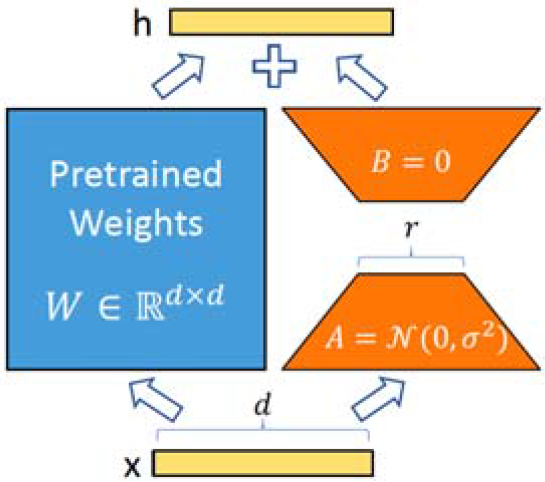
Parametrization of LoRA. Only A and B are trained. [31]

### 2.5. Classification

In the feature engineering approach, a Linear SVC was trained. When finetuning discriminative LLMs, either an MLP or a linear classifier was applied. Similarly, a linear layer was used for classification with generative LLMs. In the experiments, we also tested using generative LLMs only as the feature extractor and trained a Linear SVC for classification. In this alternative approach, which required significantly less computational resources, generative LLMs were frozen, used to encode CC texts, and the hidden states of the “Extract” token were extracted as dense representation. A Linear SVC was then trained in the similar way as in the feature engineering approach. This was to demonstrate LLMs’ native ability to understand and represent medical texts for the downstream task.

### 2.6. Optimisation

#### 2.6.1. Class weights

We also observed severe data balance in the corpora. The data imbalance ratio of GOUT-CC-2019 is (70 + 9) / 118 = 0.6695, while the imbalance ratio of GOUT-CC-2020 is (25 + 7) / 232 = 0.1379. Our first method to handle data imbalance was class weights [33], which were set according to the relative sizes of each class as in Eq. (1),

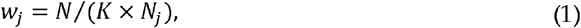

where *w*_*j*_ is the weight for the *j*-th class, *K* is the total number of classes, *N* is the total number of samples, and *N*_*j*_ is the number of samples of the *j*-th class [34].

#### 2.6.2. Oversampling

However, class weighting in Eq. (1) did not help improve the performances on GOUT-CC-2020 much, which is 5 times more imbalanced than GOUT-CC-2019. Although the discriminative LLMs performed strongly in our experiments, they were extremely sensitive to this severe data imbalance. Therefore, we performed random over sampling on GOUT-CC-2020. The positive samples in the training split, including GF-POS and GF-UNK combined, were randomly duplicated to match the size of GF-NEG.

The second approach we used to oversample the minority class was Synthetic Minority Over-sampling Technique (SMOTE) [35]. SMOTE generates synthetic examples of then minority class by interpolating the feature space of the existing minority samples. By doing so, SMOTE effectively oversamples the minority class, thereby balancing the class distribution [35]. This approach was only implemented in the method where we used the LLMs as feature extractors and classified with the SVC.

#### 2.6.3. Focal Loss

In the context of our classification tasks, the choice of a suitable loss function plays a pivotal role in training and optimizing our models. We employed two distinct loss functions as per dataset and model requirement, namely cross-entropy loss and focal loss [36], to effectively guide the training process and address specific challenges posed by our datasets.

In instances where class imbalance persisted even after oversampling the training data, such as in the case of GOUT-CC-2020, we employed focal loss as an alternative to cross-entropy to combat class imbalance in classification tasks, as in Eq. (2).

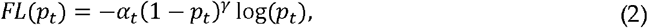

where *p*_*t*_ is the posterior probability of each target *t* (here *t*= 0 *or* 1), *α*_*t*_ ∈ [0,1] is the scaling parameter, *γ* is the focusing parameter and (1-*p*_*t*_)^*γ*^ is the modulating factor of the original cross-entropy loss [36].

## 3. Results

In this section, we meticulously analyze and compare the performances of all methods. The performance of each model was evaluated using standard metrics, including precision, recall, and Macro F1-score. We compared our results with the original algorithm proposed by Osborne et al. [3], ensuring a comprehensive assessment of the advancements achieved.

### 3.1.. Fine-tuned LLM

This subcategory encompasses results obtained by directly employing LLMs for CC classification. Table 5 shows the results.

**Table 5:** Performances of Gout Flare Detection using Fine-Tuned LLMs.

| Model | GOUT-CC-2019 |  |  | GOUT-CC-2020 |  |  |
| --- | --- | --- | --- | --- | --- | --- |
|  | Precision | Recall | F1-score | Precision | Recall | F1-score |
| **RoBERTa-Large-PM-M3-Voc** | **0.80** | **0.79** | **0.80** | 0.62 | 0.72 | 0.63 |
| **BioELECTRA** | 0.76 | 0.76 | 0.76 | 0.63 | 0.68 | 0.65 |
| **BioBART** | 0.74 | 0.73 | 0.73 | 0.65 | 0.70 | 0.67 |
| **BioGPT** | 0.62 | 0.59 | 0.60 | **0.82** | **0.88** | **0.85** |
| **BioMedLM** | 0.49 | 0.49 | 0.47 | 0.52 | 0.53 | 0.52 |

The table shows that RoBERTa-Large-PM-M3-Voc outperforms the other four models in the 2019 dataset in terms of precision, recall, and F1-score for both datasets. This suggests that this model is more effective at detecting GFs from clinical notes. Table 5 also shows that BioBERT and BioELECTRA have similar performance, while BioGPT and BioMedLM have the lowest performance among the five models.

On the 2020 dataset, the best model was by far BioGPT, outperforming others LLM competitors by large margins. Good performances were obtained due to oversampling, which improved the results from 0.67 to 0.85 macro f1 score. These results suggest that BioGPT can handle the data imbalance and the domain-specific vocabulary better than the other models, and that oversampling can boost the performance of generative LLMs for this task. On the other hand, BioMedLM did not achieve good performances, possibly due to the limitations of the LoRA adaptor, compared to BioGPT which was fully finetuned to adapt better to the special domain of gout flare CC texts.

### 3.2. Frozen LLMs as Feature Extractors

In this subcategory, we used LLMs to embed CC texts to dense feature vectors and use Linear SVC for classification. Table 6 shows the results.

**Table 6:** Performances of Gout Flare Detection using LLM Embeddings.

| Algorithm | Gout-CC-2019 |  |  | Gout-CC-2020 |  |  |
| --- | --- | --- | --- | --- | --- | --- |
|  | Precision | Recall | F1-score | Precision | Recall | F1-score |
| **SVM with BioGPT Embeddings** | 0.68 | **0.67** | **0.67** | 0.69 | **0.73** | **0.71** |
| **SVM with BioMedLM Embeddings** | 0.69 | 0.66 | 0.66 | 0.59 | 0.70 | 0.61 |
| **SVM with PMC_LLaMA_7B Embeddings** | 0.66 | 0.66 | 0.66 | 0.60 | 0.60 | 0.60 |

The table shows that SVM with BioGPT Embeddings has the best performance among the four algorithms on both datasets. It achieves an F1-score of 0.67 on Gout-CC-2019 and 0.71 on Gout-CC-2020. This indicates that this algorithm can effectively extract the relevant features from CC texts and classify them accurately.

The table also shows that SVM with BioMedLM Embeddings and SVM with PMC_Llama_7B Embeddings have similar performance, but lower than SVM with BigGPT Embeddings. They both have an F1-score of 0.66 on Gout-CC-2019 and 0.61 on Gout-CC-2020. This suggests that these algorithms are less robust and consistent in handling the variability and complexity of CC texts.

### 3.3. Sparse Text Representation

This subcategory involves performance of the traditional feature engineering approach, which generated sparse text representations using tf-idf of *n*-gram features. Contrast and compare these results against the outcomes achieved by the LLMs, providing valuable insights into the effectiveness of each approach for GF prediction. In this section we have also included the results from the original publication of Osborne et al. [3], which are shaded. All results will be discussed further in the discussion section. Table 7 shows the results.

**Table 7:** Performances of Gout Flare Detection using Sparse Text Representations.

| Algorithm | GOUT-CC-2019 |  |  | GOUT-CC-2020 |  |  |
| --- | --- | --- | --- | --- | --- | --- |
|  | Precision | Recall | F1-score | Precision | Recall | F1-score |
| **SVM with tf-idf** | 0.75 | 0.75 | 0.75 | 0.82 | 0.74 | 0.77 |
| **NAIVE-GF** | 0.23 | 1.00 | 0.38 | 0.28 | 0.56 | 0.37 |
| **SIMPLE-GF** | 0.44 | 0.84 | 0.58 | 0.37 | 0.40 | 0.38 |
| **BERT-GF** | 0.71 | 0.48 | 0.56 | 0.79 | 0.47 | 0.57 |

## 4. Discussion

### 4.1. Comparative Analysis

The following table compares the results acquired from this study, with the results obtained from the paper by Osborne et al. As shown in Table 8, RoBERTa was the best performing model on the GOUT-CC-2019-CORPUS dataset followed by BioELECTRA, showcasing the superiority of discriminative LLMs in classification tasks. The SVM with BioGPT embedding and tf-idf also performed well in relation to the other models. In the GOUT-CC-2020-CORPUS dataset the best was BioGPT which outperformed all the discriminative LLMs. This model responded very well to the fine tuning and oversampling. This result was still outperformed by SVM with tf-idf features. All our models outperformed the models used in the study by Osborne et al. (in grey) in both datasets. Overall, RoBERTa-Large-PM-M3-Voc, BioGPT and tf-idf on *n*-grams were more robust models across datasets, particularly the latter. In addition, BioGPT was a more robust feature extractor when model parameters were frozen. Finally, a promising future direction to employ the strengths of different classifier to achieve better recall while at the meantime keeping a better balance for precision.

**Table 8:** Comparing the Performances of All Gout Flare Detection Methods.

| Algorithm | GOUT-CC-2019 |  |  | GOUT-CC-2020 |  |  |
| --- | --- | --- | --- | --- | --- | --- |
|  | Precision | Recall | F1-score | Precision | Recall | F1-score |
| **RoBERTa-Large-PM-M3-Voc** | 0.80 | 0.79 | **0.80\*** | 0.62 | 0.72 | 0.63 |
| **BioELECTRA** | 0.76 | 0.76 | 0.76 | 0.63 | 0.68 | 0.65 |
| **BioBART** | 0.74 | 0.73 | 0.73 | 0.65 | 0.70 | 0.67 |
| **BioGPT** | 0.62 | 0.59 | 0.60 | **0.82** | **0.88** | **0.85** |
| **BioMedLM** | 0.49 | 0.49 | 0.47 | 0.52 | 0.53 | 0.52 |
| **SVM with BioGPT Embeddings** | 0.68 | **0.67** | **0.67** | 0.69 | 0.73 | 0.71 |
| **SVM with BioMedLM Embeddings** | 0.69 | 0.66 | 0.66 | 0.59 | 0.70 | 0.61 |
| **SVM with PMC_LLaMA_7B Embeddings** | 0.66 | 0.66 | 0.66 | 0.60 | 0.60 | 0.60 |
| **SVM with tf-idf** | 0.75 | 0.75 | 0.75 | 0.82 | 0.74 | 0.77 |
| **NAIVE-GF** | 0.23 | 1.00 | 0.38 | 0.28 | 0.56 | 0.37 |
| **SIMPLE-GF** | 0.44 | 0.84 | 0.58 | 0.37 | 0.40 | 0.38 |
| **BERT-GF** | 0.71 | 0.48 | 0.56 | 0.79 | 0.47 | 0.57 |

### 4.2. Potential and limitations

The best performance on these datasets was achieved by RoBERTa-large-PM-M3-Voc, which outperformed other LLMs and traditional machine learning algorithms. This suggests that RoBERTa-Large-PM-M3-Voc can effectively capture the semantic features of CC texts and distinguish between GF and non-flares. However, the results also show that there is still a large gap between the performance of LLMs and the desired accuracy for GF detection.

Furthermore, the results also indicate that some models have a bias towards the negative class, which may affect their ability to predict the positive label. Therefore, more research is needed to address these challenges and improve the performance of LLMs for GF detection. One of the main challenges is the nature of the dataset. All the chief complaints contain the keyword “gout” and most of them did not contain any clear indicator of gout flare. This makes it difficult for the models to learn the subtle differences between gout flares and non-flares. Upon analysing the predict column of our test set (which contains the prediction of the human annotators based solely on the CC) we found that this is a challenging problem even for professional rheumatologists which achieved less than 50% accuracy in our test set.

Although the performance on GOUT-CC-2020-CORPUS was not as good as GOUT-CC-2019-CORPUS, it’s still an improvement compared to the baseline. We acknowledge that the dataset is challenging due to its data imbalance and small size, which contributed to the performance decline. Our approaches to tackling the data imbalance did improve the performance but future work is still required to tackling this issue. One potential direction is the use of semi-supervised learning do deal with the low number of annotated CC’s and another is to encourage the medical community to share or annotate more data to create high-quality datasets.

### 4.3. Future Directions

Some improvements can be done to enhance the results obtained in this research:

#### Full Fine-Tuning and Distributed Computing

While parameter-efficient fine-tuning, specifically LoRA, was applied in this study due to hardware constraints and the models’ size, pursuing full fine-tuning would enhance the results of the models. Implementing distributed computing is necessary to apply full fine tuning, due to the very large size of the models this process requires distributing the model load across different GPUs to perform the calculations. This strategy would enable more comprehensive fine-tuning, potentially leading to an increase in model performance.

#### Enhanced Dataset Quality and Size

with such a limited number of samples the model cannot be properly trained, validated and tested. To address this more samples must be acquired or whole new datasets to test the models effectively.

#### Ensemble Learning for Enhanced Embeddings

A promising route is the utilization of deep learning models to create an ensemble that enhances embeddings before their application in text classification. This strategy could potentially enhance the information captured by the embeddings, thereby leading to improved classification outcomes.

#### Task-specific continuous pre-training

Another possible direction is to use unsupervised learning to continuously pre-train the LLMs on the task-specific data, i.e., the chief complaint texts. This could help the models to adapt to the domain and the vocabulary, and to tackle the particular write styles of keeping CC notes in the task.

## 5. Conclusions

Overall, this study highlighted the potential of generative LLMs for classification tasks, achieving results comparable to the discriminative models. Additionally, the models also have shown potential as feature extractors for classification tasks even without fine tuning, due to their ability to understand contextual information and produce contextual rich embeddings. Despite the results between the two types of models being comparable, the computational requirements to perform the same task is much greater using the generative LLMs employed in this study. Similar or superior results can be obtained using much smaller discriminative models. Still, this research highlights the importance of using the domain specific variants of the models when the text contains specialized and out of word vocabulary. Our results are important because they demonstrate the feasibility and effectiveness of using generative LLMs for gout flare detection from chief complaints, which is a novel and challenging task that can benefit both clinical practice and research. Furthermore, our approaches can potentially improve the quality of care for gout patients, a large portion of them could now receive proper and in-time follow-up after an ED visit.

## Author Contributions

Conceptualization, X. Jiang, and A. Daneshkakh; methodology, X. Jiang, L.L. Oliveira, A.N., Babu, P. Karajagi, and A. Daneshkakh; software, L.L. Oliveira, A.N., Babu, P. Karajagi, and X. Jiang; validation, L.L. Oliveira, and X. Jiang; investigation, L.L. Oliveira, A.N., Babu, P. Karajagi, and X. Jiang; resources, X. Jiang.; data curation, L.L. Oliveira; writing—original draft preparation, L.L. Oliveira, and X. Jiang; writing—review and editing, L.L. Oliveira, X. Jiang, and A. Daneshkakh; supervision, X. Jiang, and A. Daneshkakh; project administration, X. Jiang. All authors have read and agreed to the published version of the manuscript.

## Data Availability Statement

The dataset the current paper used is a public dataset, which is available through PhysioNet at https://doi.org/10.13026/96v3-dw72.

## Disclaimer/Publisher’s Note

The statements, opinions and data contained in all publications are solely those of the individual author(s) and contributor(s) and not of MDPI and/or the editor(s). MDPI and/or the editor(s) disclaim responsibility for any injury to people or property resulting from any ideas, methods, instructions or products referred to in the content.

## Footnotes

1 https://scikit-learn.org/

2 https://huggingface.co/Sedigh/RoBERTa-large-PM-M3-Voc

3 https://github.com/kamalkraj/BioELECTRA

4 https://github.com/GanjinZero/BioBART

5 https://www.nature.com/articles/sdata201635

6 https://huggingface.co/docs/transformers/model_doc/biogpt

7 https://github.com/stanford-crfm/BioMedLM

8 https://github.com/chaoyi-wu/PMC-LLaMA

## References

1. Chen, X.M.; Yokose, C.; Rai, S.K.; Pillinger, M.H.; Choi, H.K. Contemporary Prevalence of Gout and Hyperuricemia in the United States and Decadal Trends: The National Health and Nutrition Examination Survey, 2007-2016. Arthritis Rheumatol 2019, 71(6), 991–999. doi:10.1002/art.40807.

2. Singh, J.A.; Yu, S. Time Trends, Predictors, and Outcome of Emergency Department Use for Gout: A Nationwide US Study. J Rheumatol 2016, 43(8), 1581–1588. doi:10.3899/jrheum.151419.

3. Osborne, J.D.; Booth, J.S.; O’Leary, T.; et al. Identification of Gout Flares in Chief Complaint Text Using Natural Language Processing. AMIA Annu Symp Proc. 2020, 973–982.

4. Hossain, E.; Rana, R.; Higgins, N.; Soar, J.; Barua, P.D.; Pisani, A.R.; Turner, K. Natural Language Processing in Electronic Health Records in relation to healthcare decision-making: A systematic review. Comput Biol Med 2023, 155, 106649. doi: 10.1016/j.compbiomed.2023.106649.

5. Zheng, C.; Rashid, N.; Wu, Y.; et al. Using Natural Language Processing and Machine Learning to Identify Gout Flares From Electronic Clinical Notes. Arthritis Care Res 2014, 66(11), 1740–1748. doi: 10.1002/acr.22324.

6. Devlin, J.; Chang, M.W.; Lee, K.; Toutanova, K. BERT: Pre-training of Deep Bidirectional Transformers for Language Understanding. In Proceedings of the 2019 Conference of the North American Chapter of the Association for Computational Linguistics: Human Language Technologies, (NAACL-HLT’2019), 4171–4186, Minneapolis, MN, USA, 2^nd^-7^th^ June 2019. doi: 10.18653/v1/N19-1423.

7. Brown, T.; Mann, B.; Ryder, N.; et al. Language Models are Few-Shot Learners. In Advances in Neural Information Processing Systems; Larochelle, H., Ranzato, M., Hadsell, R., Balcan, M.F., Lin, H., Eds.; Curran Associates Inc., 2020; Volume 33., pp. 1877–1901. https://papers.nips.cc/paper/2020/hash/1457c0d6bfcb4967418bfb8ac142f64a-Abstract.html.

8. Xu, B.; Gil-Jardiné, C.; Thiessard, F.; Tellier, E.; Avalos, M.; Lagarde, E. Pre-training A Neural Language Model Improves The Sample Efficiency of an Emergency Room Classification Model. In Proceedings of The Thirty-Third International FLAIRS Conference (FLAIRS-33), North Miami Beach, Florida, USA, 17-20 May 2020. https://aaai.org/papers/264-flairs-2020-18444/.

9. Veladas, R.; Yang, H.; Quaresma, P.; et al. Aiding Clinical Triage with Text Classification. In Progress in Artificial Intelligence, Lecture Notes in Computer Science; Marreiros, G., Melo, F.S., Lau, N., Lopes Cardoso, H., Reis, L.P., Eds.; Springer International Publishing; 2021; Vol 12981, pp. 83–96. doi: 10.1007/978-3-030-86230-5_7.

10. Ding, L.; Zhang, Z.; Liu, H.; Li, J.; Yu, G. Automatic Keyphrase Extraction from Scientific Chinese Medical Abstracts Based on Character-Level Sequence Labeling. J Data Inf Sci 2021, 6(3), 35–57. doi: 10.2478/jdis-2021-0013.

11. Ding, L.; Zhang, Z.; Zhao, Y. Bert-Based Chinese Medical Keyphrase Extraction Model Enhanced with External Features. In: Towards Open and Trustworthy Digital Societies, Lecture Notes in Computer Science; Ke, H.R., Lee, C.S., Sugiyama, K., Eds.; Springer International Publishing; 2021; Volume 13133, pp. 167–176. doi: 10.1007/978-3-030-91669-5_14.

12. Han, L.; Erofeev, G.; Sorokina, I.; Gladkoff, S.; Nenadic, G.; Investigating Massive Multilingual Pre-Trained Machine Translation Models for Clinical Domain via Transfer Learning. In Proceedings of the 5th Clinical Natural Language Processing Workshop (ClinicalNLP’2019), 31–40, Minneapolis, MN, USA, 7^th^ June 2019. doi: 10.18653/v1/2023.clinicalnlp-1.5.

13. Tang, L.; Sun, Z.; Idnay, B.; et al. Evaluating Large Language Models on Medical Evidence Summarization. npj Digit. Med. 2003, 6, Article No. 158. doi: 10.1038/s41746-023-00896-7.

14. Osborne, J. D., O’Leary, T., Mudano, A., Booth, J., Rosas, G., Peramsetty, G. S., Knighton, A., Foster, J., Saag, K., & Danila, M. I. Gout Emergency Department Chief Complaint Corpora (version 1.0). PhysioNet, 2020. doi: 10.13026/96v3-dw72.

15. Manning, C.D.; Raghavan, P.; Schütze, H. Introduction to Information Retrieval; Cambridge University Press: Cambridge, USA, 2008.

16. Vaswani, A.; Shazeer, N.; Parmar, N.; et al. Attention is All you Need. In Advances in Neural Information Processing Systems; Guyon, I., Luxburg, U.V., Bengio, S., et al., Eds.; Curran Associates, Inc., 2017; Volume 30, pp. 5998–6008. https://papers.nips.cc/paper_files/paper/2017/hash/3f5ee243547dee91fbd053c1c4a845aa-Abstract.html.

17. Kanakarajan, K.R.; Kundumani, B.; Sankarasubbu, M. BioELECTRA: Pretrained Biomedical Text Encoder using Discriminators. In Proceedings of the 20th Workshop on Biomedical Language Processing (BioNLP’2021), 143–154. Online, 16 August 2021. doi: 10.18653/v1/2021.bionlp-1.16.

18. Yuan, H.; Yuan, Z.; Gan, R.; Zhang, J.; Xie, Y.; Yu, S. BioBART: Pretraining and Evaluation of A Biomedical Generative Language Model. In Proceedings of the 21st Workshop on Biomedical Language Processing (BioNLP’2022), 97–109. Dublin, Ireland, w6th May 2022. doi: 10.18653/v1/2022.bionlp-1.9.

19. Liu, Y.; Ott, M.; Goyal, N.; et al. RoBERTa: A Robustly Optimized BERT Pretraining Approach. http://arxiv.org/abs/1907.11692.

20. Clark, K.; Luong, M.T.; Le, Q.V.; Manning, C.D. ELECTRA: Pre-training Text Encoders as Discriminators Rather Than Generators. In Proceedings of the Eighteenth International Conference on Learning Representations (ICLR’2020). Online, 27^th^-30^th^ April 2020. https://openreview.net/forum?id=r1xMH1BtvB.

21. Lewis, M.; Liu, Y.; Goyal, N.; et al. BART: Denoising Sequence-to-Sequence Pre-training for Natural Language Generation, Translation, and Comprehension. In Proceedings of the 58th Annual Meeting of the Association for Computational Linguistics (ACL’2020), 7871–7880. Online, 5^th^-10^th^ July 2020. doi: 10.18653/v1/2020.acl-main.703.

22. Johnson, A.E.W.; Pollard, T.J.; Shen, L.; et al. MIMIC-III, a freely accessible critical care database. Sci Data 2016, 3(1): 160035. doi: 10.1038/sdata.2016.35.

23. Luo, R.; Sun, L.; Xia, Y.; et al. BioGPT: generative pre-trained transformer for biomedical text generation and mining. Brief Bioinform 2022, 23(6), bbac409. doi: 10.1093/bib/bbac409.

24. Wu, C.; Lin, W.; Zhang, X.; Zhang, Y.; Wang, Y.; Xie, W. PMC-LLaMA: Towards Building Open-source Language Models for Medicine. Preprint, 2023. https://arxiv.org/abs/2304.14454.

25. Radford, A.; Wu, J.; Child, R.; Luan, D.; Amodei, D.; Sutskever, I. Language Models are Unsupervised Multitask Learners. Technical Report, 2018. https://d4mucfpksywv.cloudfront.net/better-language-models/language-models.pdf.

26. Touvron, H.; Lavril, T.; Izacard, G.; et al. LLaMA: Open and Efficient Foundation Language Models. Preprint, 2023. https://arxiv.org/abs/2302.13971.

27. Touvron, H.; Martin, L.; Stone, K.; et al. Llama 2: Open Foundation and Fine-Tuned Chat Models. Preprint, 2023. http://arxiv.org/abs/2307.09288.

28. Gao, L.; Biderman, S.; Black, S.; et al. The Pile: An 800GB Dataset of Diverse Text for Language Modeling. Preprint, 2021. http://arxiv.org/abs/2101.00027.

29. Lo, K.; Wang, L.L.; Neumann, M.; Kinney, R.; Weld, D. S2ORC: The Semantic Scholar Open Research Corpus. In Proceedings of the 58th Annual Meeting of the Association for Computational Linguistics (ACL’2020), 4969–4983. Online, 5^th^-10^th^ July 2020. doi: 10.18653/v1/2020.acl-main.447.

30. Loshchilov, I., Hutter, F. Decoupled Weight Decay Regularization. In Proceedings of the Seventh International Conference on Learning Representations (ICLR’2019). New Orleans, USA, 6^th^-9^th^ May 2019. https://openreview.net/pdf?id=Bkg6RiCqY7.

31. Hu, E.J.; Shen, Y.; Wallis, P.; et al. LoRA: Low-Rank Adaptation of Large Language Models. In Proceedings of the Ninth International Conference on Learning Representations (ICLR’2021). Online, 3^rd^-7^th^May 2021. https://openreview.net/forum?id=nZeVKeeFYf9.

32. Aghajanyan, A.; Zettlemoyer, L.; Gupta, S. Intrinsic Dimensionality Explains the Effectiveness of Language Model Fine-Tuning. In Proceedings of the 59th Annual Meeting of the Association for Computational Linguistics and the 11th International Joint Conference on Natural Language Processing (ACL-IJCNLP’2020), 7319–7328. Online, 1^st^-6^th^ August 2020. Doi: 10.18653/v1/2021.acl-long.568.

33. He, J.; Cheng, X. Weighting Methods for Rare Event Identification From Imbalanced Datasets. Front Big Data 2021, Volume 4, Article 715320. doi: 10.3389/fdata.2021.715320.

34. Singh, K. How to Improve Class Imbalance using Class Weights in Machine Learning? Analytics Vidhya. Published October 6, 2020. https://www.analyticsvidhya.com/blog/2020/10/improve-class-imbalance-class-weights. Accessed on 29th January 2024.

35. Chawla, N.V.; Bowyer, K.W.; Hall, L.O.; Kegelmeyer, W.P. SMOTE: Synthetic Minority Over-sampling Technique. J Artif Intell Res 2002, 16, 321–357. doi: 10.1613/jair.953.

36. Lin, T.Y.; Goyal, P.; Girshick, R.; He, K.; Dollar, P. Focal Loss for Dense Object Detection. IEEE Trans Pattern Anal Mach Intell 2020, 42(2), 318–327. doi: 10.1109/TPAMI.2018.2858826.

